# Secular trends in age at menarche and associated determinants in the Valencian population (Spain)

**DOI:** 10.64898/2026.04.29.26351926

**Authors:** Andrea Beneito, Blanca Sarzo, Raul Beneyto, Reem Abumallouh, Natalia Marin, Oihane Alvarez, Ana Molina-Barceló, Mercedes Vanaclocha-Espí, Carmen Freire, Ferran Ballester, Ana Esplugues, Maria-Jose Lopez-Espinosa

## Abstract

**Background:** Menarche is a critical developmental milestone, with earlier onset associated with adverse long-term health consequences. Despite a reported global decline in age at menarche over the last century, this trend and its determinants remain insufficiently studied in Spain.

**Objective:** To assess secular trends in age at menarche and its determinants in the Valencian Community, Spain.

**Methods:** This population-based study included 417,260 participants born between 1931 and 2008. First, secular trends in age at menarche were assessed using time-series models across 5-year birth cohorts for the overall population. Then, participants were categorized as either women (born 1931–1985) or girls (born 1990–2008), and Bayesian linear regression models were fitted for each group, adjusting for birth cohort and continent of birth in all models, and additionally for educational level in women and body mass index (BMI) in girls.

**Results:** Mean age at menarche decreased by 1.9 years, from 13.1 to 11.1, between the 1931–1935 and 2006–2008 birth cohorts, with a steeper decline after 1975. Compared to Europeans, women born in South/Central America (β[95% CI]: 0.33[0.30, 0.36] years) and Africa (0.52[0.45, 0.58] years) experienced later menarche, while girls from South/Central America experienced earlier onset (−0.18[−0.28, −0.09] years). In girls, lower BMI was associated with later menarche (0.96[0.74, 1.18] years) and higher BMI with earlier onset (−0.53[−0.57, −0.48] years).

**Conclusion:** There was a marked decline in age at menarche in the Valencian Community, with no evidence of leveling off. Key determinants included continent of birth (with cohort-specific effects) and BMI.

## 1. Introduction

Menarche, defined as the first menstrual period, is a fundamental milestone in female pubertal development that marks the onset of reproductive capacity[1]. It is a late pubertal indicator that occurs at a mean age ± standard deviation (SD) of 12.4±2.5 years, with a typical range of 10 to 16 years[1]. Its timing is regulated by the endocrine–reproductive system[1] and is primarily driven by rising estrogen levels[2,3]. Beyond biological pubertal processes, menarche involves emotional, psychosocial, cognitive, and physical changes that mark the transition to adolescence[3,4].

At the population level, menarche timing is of public health importance, given the well-documented associations between earlier menarche and adverse long-term health consequences, including increased risks of hormone-related cancers[2,5–8], cardiovascular and metabolic diseases[9–11], and mental health disorders[12]. In this context, a substantial body of literature has documented a consistent global secular trend toward earlier menarche. Specifically, historical data from Western countries show a decline from ∼17 years in the early 19th century to ∼13 years by the mid-20th century[13,14]. In low- and middle-income countries, where ongoing socioeconomic transitions and improvements in nutrition and healthcare are occurring, the age at menarche is also declining[15–17].

Taken together, these temporal trends suggest that menarche timing is influenced by a complex interplay of biological, nutritional, environmental, and socioeconomic factors, with genetic predisposition providing the biological framework[18,19] and the other factors shaping its expression across the life course[20,21]. Historically, improvements in living standards (particularly in healthcare, socioeconomic conditions, and nutrition) have contributed to earlier menarche[22], whereas unfavorable conditions have been associated with delays[21–23].

Although the secular decline in menarche age is well documented globally, evidence from Spain remains limited. The available Spanish studies[13,24–26] are scarce and outdated, preventing a comprehensive assessment of the magnitude of the secular decline in age at menarche and of whether the trend has continued, stabilized, or reversed over time. Moreover, the underlying factors influencing patterns of menarcheal timing remain poorly understood in the Spanish context, further highlighting the need for updated, population-based evidence. To address these gaps, this study examines the secular trend in age at menarche and its determinants among 417,260 women and girls from the Valencian Community (Spain) born between 1931 and 2008.

## 2. Methods

### 2.1. Study population

This study used four data sources from the Valencian Community (Spain): two population databases and two cohort studies. The first population-based database comprised women participating in the Valencian Breast Cancer Screening Program (BCSP)[27]. It included 398,074 women (born 1931–1974) with data recorded between 1992 and 2019 at ages 45–65 years. The second population-based database included public healthcare records from the Ambulatory Care Information System of the Valencian Health Agency (SIA-GAIA)[28], comprising 20,381 girls with data from primary care visits (born 1990–2008; data collected 2004–2010 at ages 8–16). Regarding the cohorts, the INMA-Valencia cohort[29] recruited pregnant women from the Valencia area in 2003–2005, with mothers and offspring followed up prospectively for over 20 years. This database included 132 INMA girls (born 2004–2006; surveys conducted 2013–2021 at ages 9–16 years) and 282 INMA mothers (born 1961–1985; surveyed 2019–2021 at ages 34–57 years). The second cohort, PAPILONG[30], comprised socially and economically vulnerable women (i.e., sex workers, NGO-supported women, and those living in extreme poverty) residing in Valencia province, including 131 women (born 1952–1983; data collected 2021–2023 at ages 39–69 years).

A total of 419,000 participants were identified across all databases (Fig. S1, Tables S1–S2). Of these, 1,740 (0.42%) were excluded due to missing data on age at menarche or implausible values (<8 or >18 years). The remaining sample included 417,260 participants born between 1931 and 2008 and was used to assess temporal trends in age at menarche.

Then, analyses of determinants were conducted in 45.1% of the population (Fig. S1, Table S3). To this end, participants with incomplete data on the determinants under study were excluded, as were those belonging to the 1931–1940, 1976–1990, and 2006–2008 birth cohorts (owing to limited availability of determinant data, with the most recent cohort being truncated due to the lack of data beyond 2008). In addition, participants born in North America, Oceania, Asia (for both analyses), and Africa (girls only) were excluded due to small sample sizes. The final sample was stratified into women (born 1941–1975; N=177,025) and girls (born 1991–2005; N=11,867) to conduct the study of the associated factors, as differences in available determinants precluded joint analyses.

All study protocols, including two secondary population-based registries and two datasets collected by the research team, were approved by the DGSP-CSISP Ethics Committee from Valencia (references numbers: 20180112/03; 20190329/3; 20190301/09/1; and 20210604/10/01). Secondary data were provided anonymized, while investigator-collected data were anonymized prior to analysis following written informed consent from participants or legal guardians (for minors). All procedures were conducted in accordance with the Declaration of Helsinki.

### 2.2. Age at menarche

Age at menarche was retrospectively self-reported in all adult databases, either during routine clinical visits (BCSP) or through interviewer-administered questionnaires conducted during research surveys (INMA mothers and PAPILONG). For child databases, age at menarche was collected through routine clinical visits (SIA-GAIA) or follow-up visits (INMA girls), based on reports from parents and/or the girls themselves. In INMA girls, when multiple or discrepant reports were available, predefined harmonization criteria were applied by averaging parent and girl reports from the same visit or by retaining the report closest to the event[31].

### 2.3. Determinants of age at menarche

As data were pooled from multiple sources, analyses were restricted to covariates available in the two large population-based databases (i.e., year of birth, country of birth, educational level, and body mass index [BMI]). Specifically, year of birth and country of birth (grouped by continent) were available for the whole population. Educational level was available for women in BCSP, INMA mothers, and PAPILONG, and categorized as primary, secondary, or university. BMI near menarche was available only for SIA-GAIA and INMA girls. BMI z-scores were calculated from weight and height using the WHO 2006 Child Growth Standards[32] and classified as low (≤–2 SD), normal (>–2 to ≤1 SD), or high (>1 SD).

### 2.4. Statistical analysis

First, a time-series model was applied to characterize the overall pattern of age at menarche and its secular trend in the full sample of participants (N=417,260). Although year of birth was the most precise variable available, some years were only represented by a small number of women, which limited the stability of yearly estimates. Therefore, women were grouped into 5-year birth cohorts to reduce variability. Since the second analysis was limited to a subsample with complete information on all determinants, we conducted a sensitivity analysis restricting the sample to this subset and compared the resulting curves with those from the full study population (time-series model) by overlaying them in the same graph. Stationarity of the 5-year series was assessed using the augmented Dickey–Fuller test[33]. Holt’s linear trend method[34] was used to predict the long-term trend and estimate future age at menarche.

Second, analyses of determinants were conducted in a reduced subsample. Specifically, the population was stratified into women (born 1941–1975; N=177,025) and girls (born 1991–2005; N=11,867) according to differences in available data on determinants. Educational level was only available for women, as younger participants had not yet completed their education, whereas BMI was only analyzed in younger cohorts, as available measurements were closer to menarche. Accordingly, age at menarche for women was modeled as a function of birth year, birth continent, and educational level, whereas for girls, models included birth year, birth continent, and BMI. For these analyses, Bayesian linear models were fitted using the JAGS software[35]. The final approximated posterior distributions were obtained using three chains and 3,000 iterations, discarding the first 300 as burn-in. Convergence was assessed via the Brooks–Gelman–Rubin (BGR) statistic[36]. Results were presented as posterior means (β) and 95% credible intervals (95% CI) from the marginal posterior distribution of model parameters associated with each covariate. Additionally, the posterior mean of age at menarche and its 95% CI were calculated.

Sensitivity analyses were also performed to address potential cohort overlap. In the analysis of women, 217 INMA mothers and 110 PAPILONG participants were excluded due to possible overlap with the BCSP dataset. In the analysis of girls, 112 INMA girls were excluded due to potential overlap with the SIA-GAIA dataset. Results remained materially unchanged (data not shown).

Statistical analyses were performed with R software version 4.4.2[37].

## 3. Results

### 3.1. Study population

The study comprised 417,260 participants, 91.1% of whom were born in Europe. Of these, 95.1% were women (born 1931–1985), while the remainder were girls (born 1990–2008). Among women, 22.3% had completed secondary education and 15.4% university studies. Among girls, 0.97% and 43.5% had low and high BMI, respectively (Table I).

**Table I.**
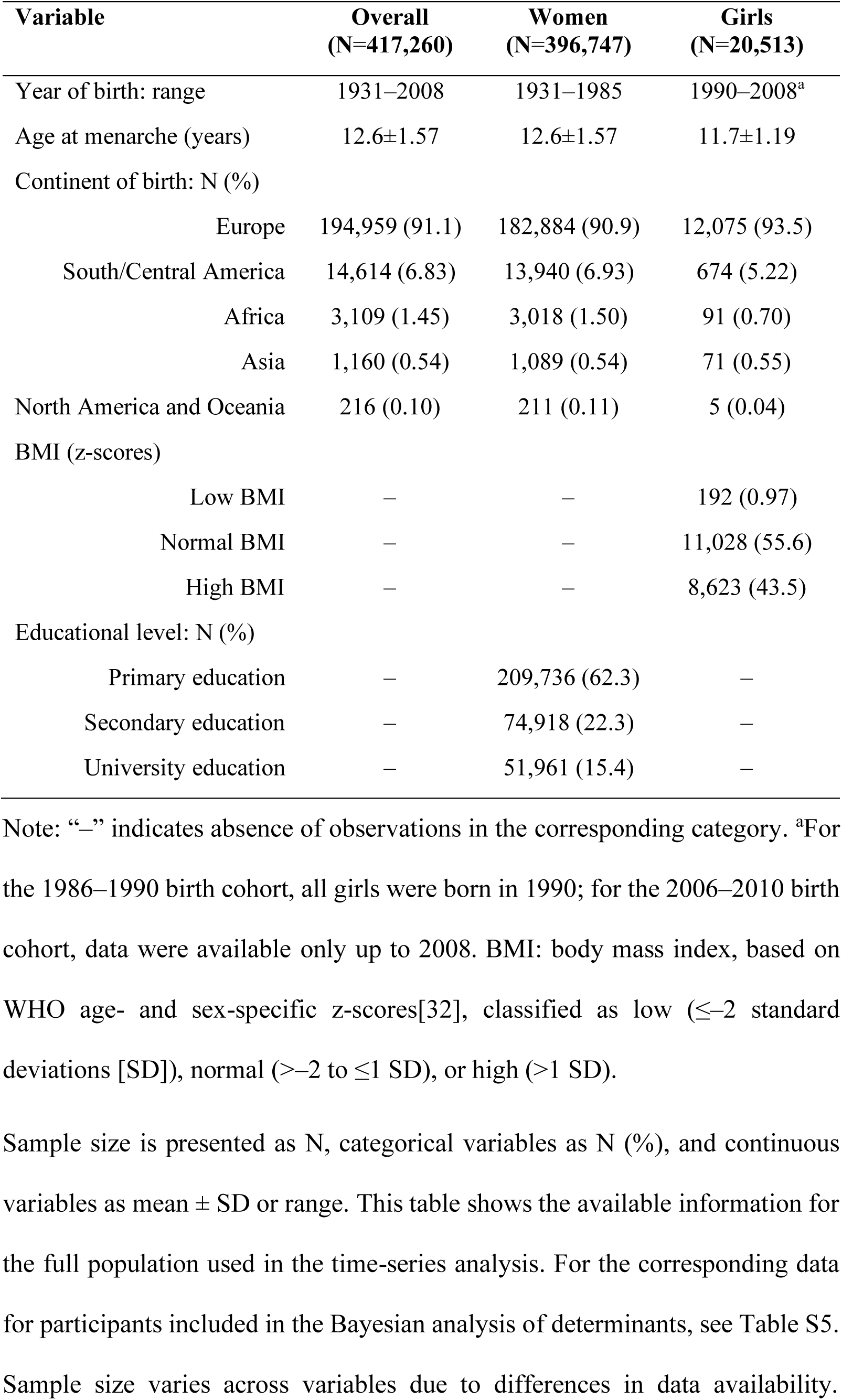
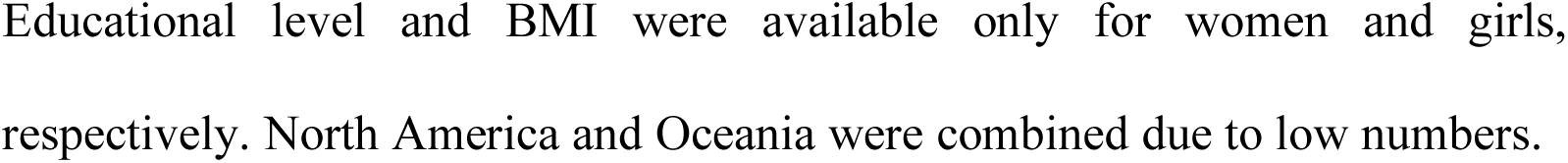
Characteristics of the study population (1931–2008)

| Variable | Overall<br>(N=417,260) | Women<br>(N=396,747) | Girls<br>(N=20,513) |
| --- | --- | --- | --- |
| Year of birth: range | 1931–2008 | 1931–1985 | 1990–2008<sup>a</sup> |
| Age at menarche (years) | 12.6±1.57 | 12.6±1.57 | 11.7±1.19 |
| Continent of birth: N (%) |  |  |  |
| Europe | 194,959 (91.1) | 182,884 (90.9) | 12,075 (93.5) |
| South/Central America | 14,614 (6.83) | 13,940 (6.93) | 674 (5.22) |
| Africa | 3,109 (1.45) | 3,018 (1.50) | 91 (0.70) |
| Asia | 1,160 (0.54) | 1,089 (0.54) | 71 (0.55) |
| North America and Oceania | 216 (0.10) | 211 (0.11) | 5 (0.04) |
| BMI (z-scores) |  |  |  |
| Low BMI | – | – | 192 (0.97) |
| Normal BMI | – | – | 11,028 (55.6) |
| High BMI | – | – | 8,623 (43.5) |
| Educational level: N (%) |  |  |  |
| Primary education | – | 209,736 (62.3) | – |
| Secondary education | – | 74,918 (22.3) | – |
| University education | – | 51,961 (15.4) | – |

Note: “–” indicates absence of observations in the corresponding category. <sup>a</sup>For the 1986–1990 birth cohort, all girls were born in 1990; for the 2006–2010 birth cohort, data were available only up to 2008. BMI: body mass index, based on WHO age- and sex-specific z-scores[32], classified as low ( $\leq -2$ standard deviations [SD]), normal ( $> -2$ to $\leq 1$ SD), or high ( $> 1$ SD).
Educational level and BMI were available only for women and girls, respectively. North America and Oceania were combined due to low numbers.

### 3.2. Secular trend of age at menarche

Overall, the mean age at menarche ± SD was 12.6±1.57 years: 12.6±1.57 years for women and 11.7±1.19 years for girls (Table I). Analyses across 5-year birth cohorts suggested a gradual decrease in age at menarche over time. The highest mean ± SD was 13.1±1.65 years between 1931–1935, and the lowest was 11.1±0.84 years in 2006–2008 (the first and last periods examined). However, mean age at menarche did not decrease linearly across birth years. There was an initial gradual decline until 1960, followed by a period of relative stability (1961–1975) and a more pronounced decrease from 1976 onwards (Fig. 1 and Table S3). Secular trends were similar in the full sample and in the subsample with determinant data (Fig. S2).

**Figure 1.**
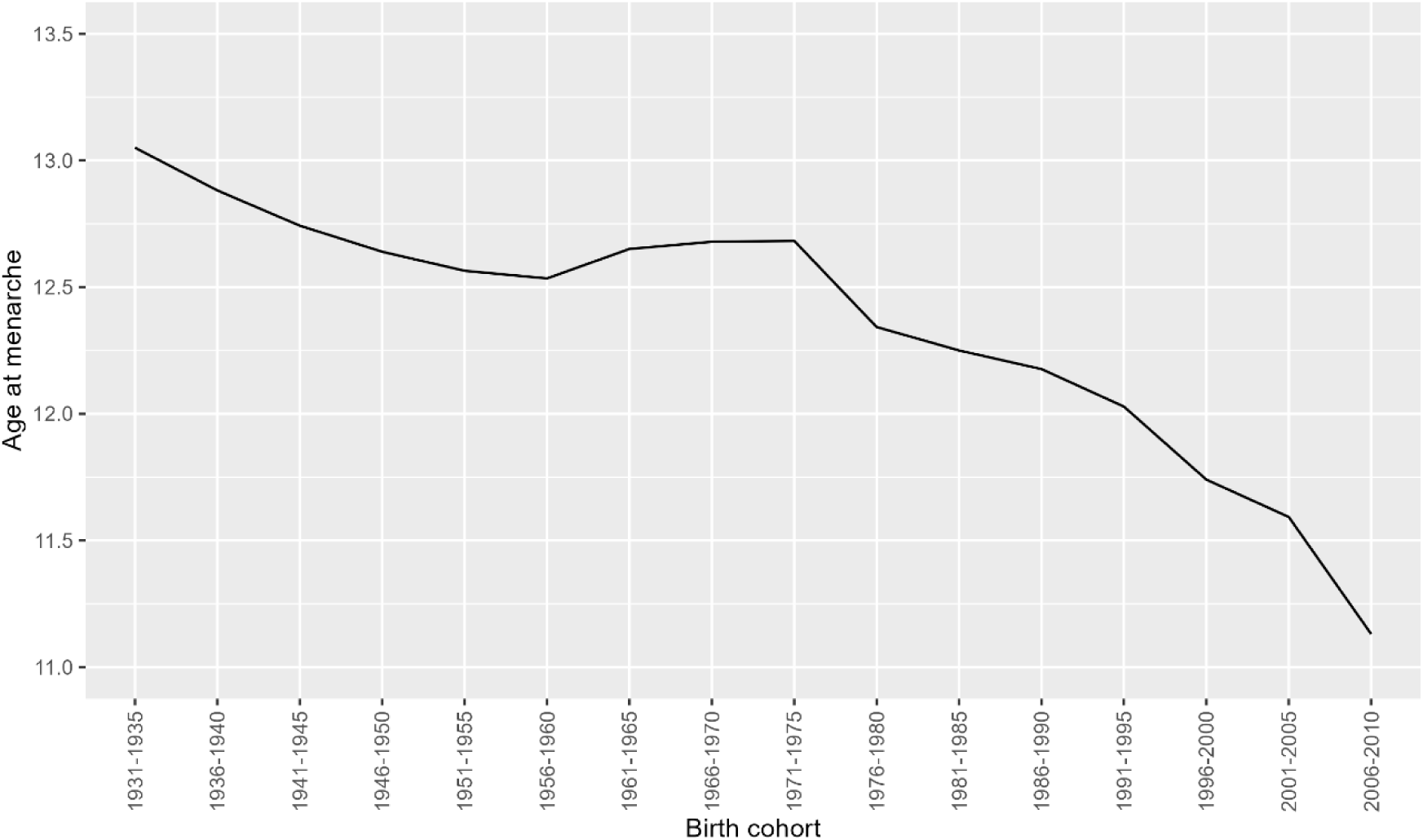
Secular trend in age at menarche by 5-year birth cohort from 1931 to 2008 (N=417,260) Note: For the most recent 5-year birth cohort (2006–2010), data availability was limited to births up to 2008.

We also made a long-term prediction of the secular trend in menarche onset. The findings indicate that it will continue to decline, with a predicted mean value (95% CI) of 9.58 (8.20, 11.0) years for the final predicted period (2031–2035) (Table S4 and Fig. S3).

### 3.3. Age at menarche and determinants

Among women (N=177,025), a slight downward trend in age at menarche was observed across birth cohorts. The posterior mean age (β [95% CI]) declined from 12.8 (12.7–12.9) to 12.6 (12.6–12.7) years for women born in 1941–1945 and 1971–1975 (Table II). Compared with women born in Europe, those born in South/Central America and Africa reported a relevant later age at menarche (0.33 [0.30, 0.36] and 0.52 [0.45, 0.58] years, respectively). Compared with women with primary education, those with secondary and university education reached menarche modestly earlier, with estimates of −0.02 (−0.03, −0.01) years and −0.01 (−0.03, 0.01) years, respectively, although the association was not relevant among university-educated women (Tables II and S5 and Fig. 2).

**Figure 2.**
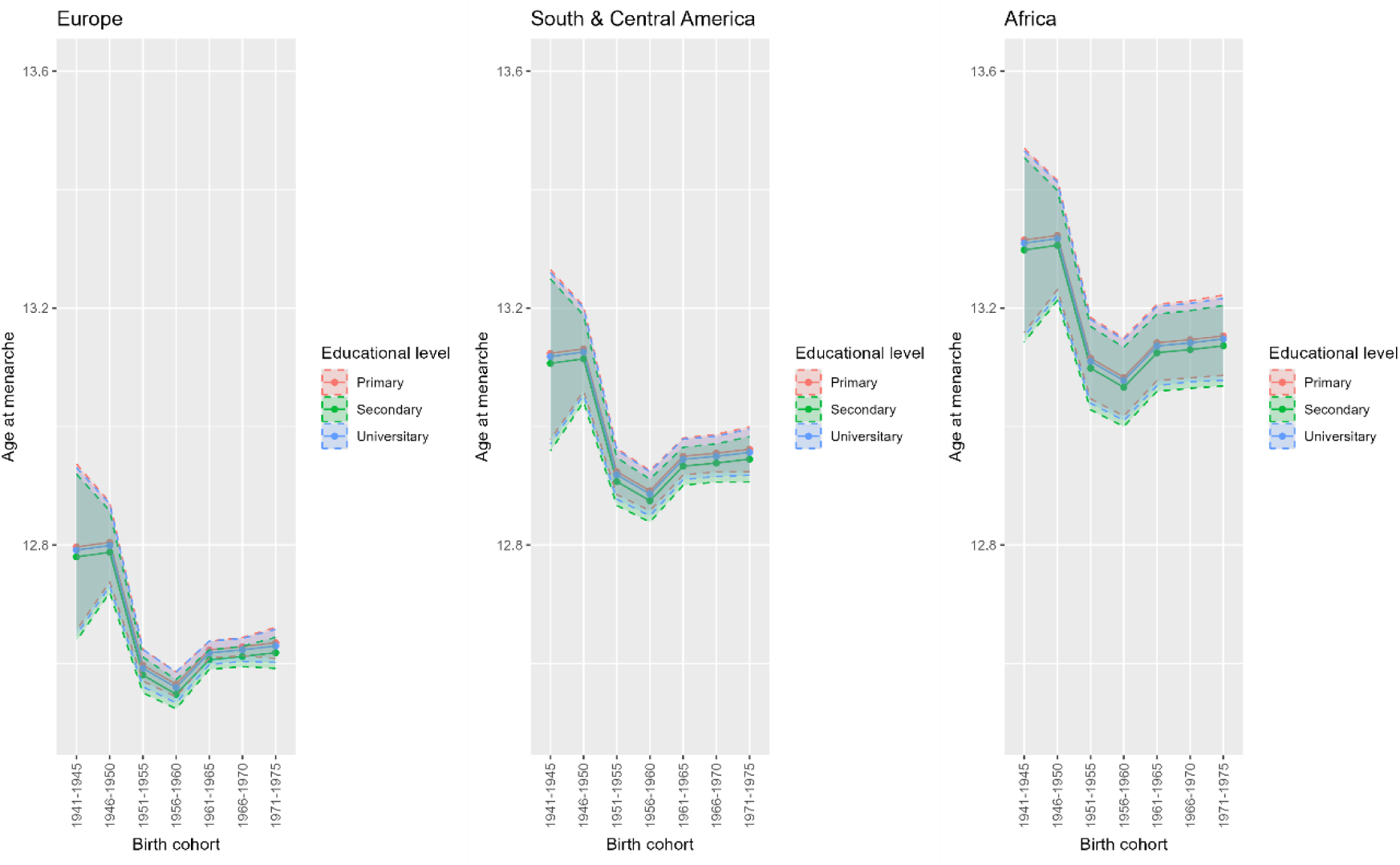
Posterior mean and 95% credible interval (95% CI) of age at menarche in women, by 5-year birth cohort, continent of birth, and educational level (N=177,025) Note: Posterior means and 95% credible intervals were estimated using Bayesian linear regression models. Results are shown for women across by 5-year birth cohort for continent of birth, and educational level.

**Table II.**
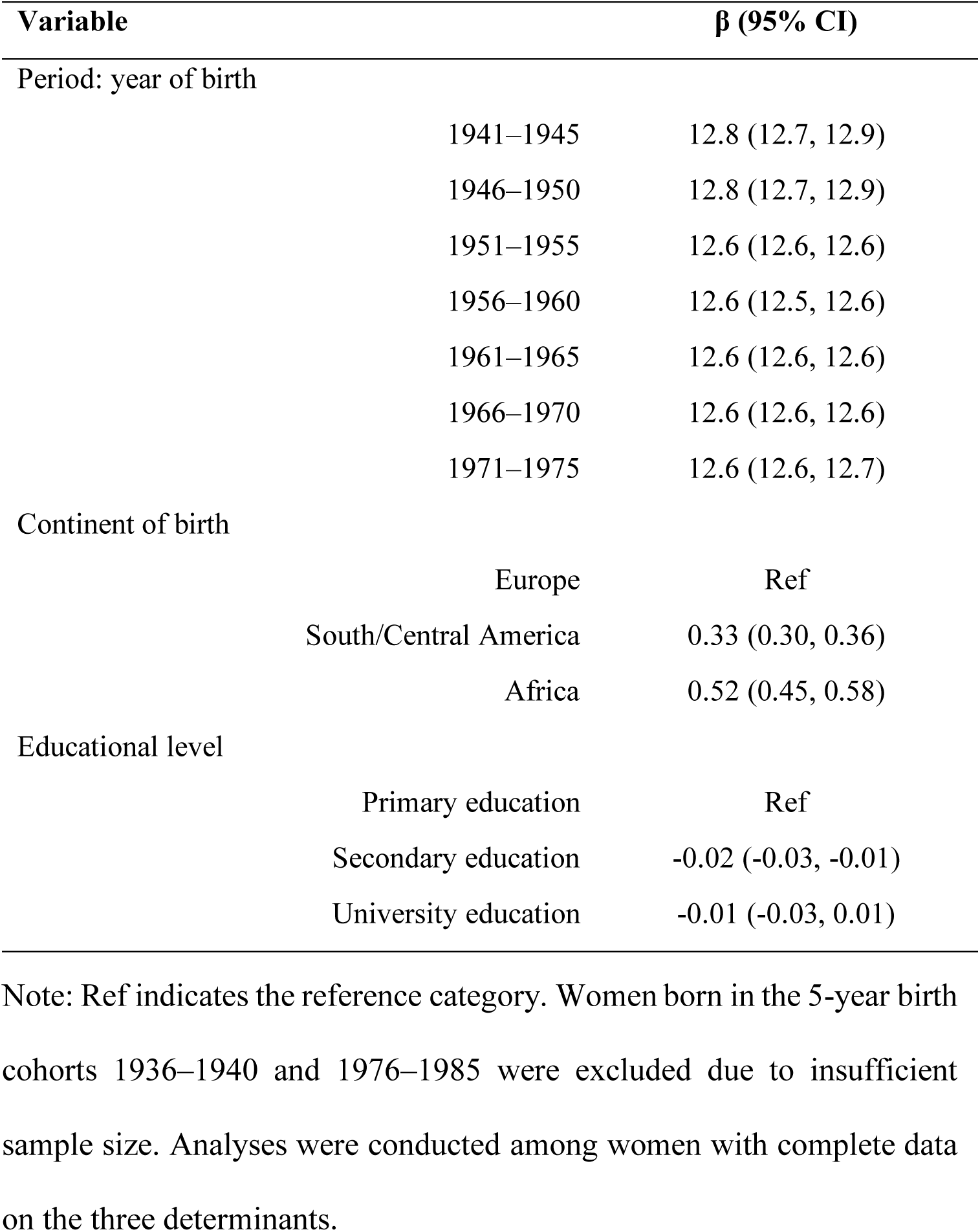
Posterior mean (β) and 95% credible interval (95% CI) for model parameters in the model for women (born 1941–1975, N=177,025)

| Variable | $\beta$ (95% CI) |
| --- | --- |
| Period: year of birth |  |
| 1941–1945 | 12.8 (12.7, 12.9) |
| 1946–1950 | 12.8 (12.7, 12.9) |
| 1951–1955 | 12.6 (12.6, 12.6) |
| 1956–1960 | 12.6 (12.5, 12.6) |
| 1961–1965 | 12.6 (12.6, 12.6) |
| 1966–1970 | 12.6 (12.6, 12.6) |
| 1971–1975 | 12.6 (12.6, 12.7) |
| Continent of birth |  |
| Europe | Ref |
| South/Central America | 0.33 (0.30, 0.36) |
| Africa | 0.52 (0.45, 0.58) |
| Educational level |  |
| Primary education | Ref |
| Secondary education | -0.02 (-0.03, -0.01) |
| University education | -0.01 (-0.03, 0.01) |

Note: Ref indicates the reference category. Women born in the 5-year birth cohorts 1936–1940 and 1976–1985 were excluded due to insufficient sample size. Analyses were conducted among women with complete data on the three determinants.

Among girls (N=11,867), those born in 1991–1995 had a slightly higher age at menarche than those born in 1996–2000 and 2001–2005 (12.2 [12.2, 12.3]; 12.0 [11.9, 12.0]; and 11.9 [11.8, 11.9] years, respectively). Girls from South/Central America had a lower age at menarche than those born in Europe (−0.18 [−0.28, −0.09] years). Compared with normal BMI, low BMI was associated with later menarche (0.96 [0.74, 1.18] years), and high BMI with earlier menarche (−0.53 [−0.57, −0.48] years) (Tables III and S5 and Fig. 3).

**Table III.** Posterior mean (β) and 95% credible interval (95% CI) for model parameters in the model for girls (born 1991–2005; N=11,867)

| Variable | $\beta$ (95% CI) |
| --- | --- |
| Period: year of birth |  |
| 1991–1995 | 12.2 (12.2, 12.3) |
| 1996–2000 | 12.0 (11.9, 12.0) |
| 2001–2005 | 11.9 (11.8, 11.9) |
| Continent of birth |  |
| Europe | Ref |
| South/Central America | -0.18 (-0.28, -0.09) |
| BMI (z-scores) |  |
| Normal BMI | Ref |
| Low BMI | 0.96 (0.74, 1.18) |
| High BMI | -0.53 (-0.57, -0.48) |

Note: BMI: body mass index, based on WHO age- and sex-specific z-scores[32], classified as low ( $\leq -2$ standard deviations [SD]), normal ( $> -2$ to $\leq 1$ SD), or high ( $> 1$ SD); Ref indicates the reference category. Girls born in the 5-year birth cohorts 1986–1990 and 2006–2008 were excluded due to insufficient sample size. No data were available for 2009–2010.

**Figure 3.**
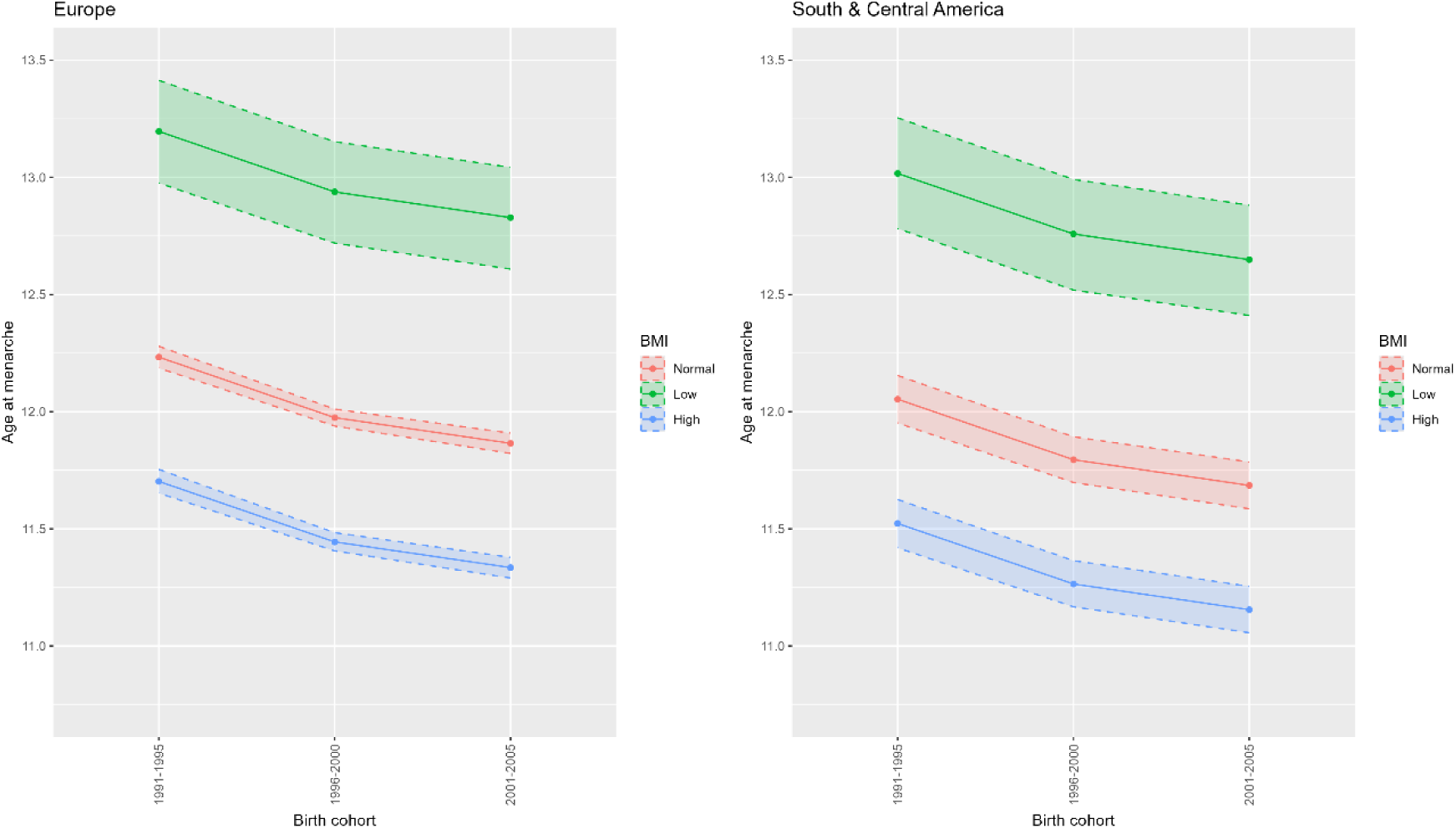
Posterior mean and 95% credible interval (95% CI) of age at menarche in girls, by 5-year birth cohort, continent of birth, and BMI (N=11,867) Note: Posterior means and 95% credible intervals were estimated using Bayesian linear regression models. Results are shown for girls across by 5-year birth cohort for continent of birth, and BMI. BMI: body mass index, based on WHO age- and sex-specific z-scores[32], classified as low (≤–2 standard deviations [SD]), normal (>–2 to ≤1 SD), or high (>1 SD).

## 4. Discussion

In our study of 417,260 participants (born 1931–2008) residing in the Valencian Community, age at menarche declined by approximately 1.9 years over seven decades, from 13.1 to 11.1 years. The association with continent of birth varied by birth group: later menarche among South/Central American and African women, and earlier onset among South/Central American girls. Additionally, higher BMI was associated with earlier age at menarche in girls. Associations with educational level were modest in magnitude and did not show a clear, statistically significant gradient across categories in women.

### 4.1. Secular trend in age of menarche

Although our findings indicate a long-term decline in the age at first menstruation, when results are broken down by 5-year birth cohort, this secular decline is steady and gradual in the early decades. Mean ages fell from 13.1 years (born 1931–1935) to 12.5 years (born 1956–1960). These results may reflect historical improvements in nutrition, living conditions, and healthcare in Spain between 1930 and 1960, although these trends were likely disrupted by the Spanish Civil War (1936–1939) and its aftermath[24,38]. This pattern is consistent with two studies conducted in other Spanish regions[24,25], which also reported a mild and progressive menarche reduction from the mid-1920s to 1960. Additionally, a similar generalized decline throughout the first half of the 20th century was reported in two studies including nine European[26] and eleven Western[13] countries, both of which included Spain. Interestingly, across these four studies, mean age at menarche (visually estimated from published figures in some cases) declined from approximately 14.8 years in the 1930s to 12.2 in the 1960s[13,24–26].

After the initial decline, age at menarche stabilized in women born between 1961 and 1975 (mean age: 12.7 years). This plateau may reflect a period in Spain without major improvements in nutrition and childhood health before increases in adiposity and lifestyle changes associated with the nutritional transition became pronounced[24,38]. Similar plateaus were observed in two Spanish studies (born 1959–1964: mean age 12.5–12.6 years[26]; and 1959–1962: 12.8–13.0 years[24]) and one Norwegian study (1955–1964: 13.2–13.2 years)[39], with a longer plateau observed in Portugal (1950–1979: 12.5–12.5 years)[40], UK (1945–1989: 12.6–12.6 years)[41], and Japan (1960–1980: 12.3–12.2 years)[42]. Some of the aforementioned data comes from visual inspection of figures in the respective articles.

From 1976 to 2008, age at menarche showed a marked overall decline in our study (12.3 to 11.1 years), despite a brief increase in the mid-1980s that should be interpreted cautiously due to the small number of available observations between 1976 and 1990 (N=137). Consistent with this decline, our projections up to 2035 suggest a continued downward trend, although the wide 95% CIs (particularly for the most distant projections) warrant cautious interpretation. The decline that resumed in cohorts born after the mid-1970s coincided with the Spanish democratic transition, a period characterized by rapid socioeconomic changes and an intensified nutritional transition[38]. This overall decrease, though more pronounced, aligns with declines reported in Portugal (born 1970–2000: 12.5–12.0 years)[40], Taiwan (1975–1989: 12.9–12.2 years)[16], and the UK (1985–1993: 12.6–12.3 years)[41]. Additionally, a multi-country study in Europe and the United States suggested that the secular decline has attenuated since the 1960s, although statistically significant decreases of 2.5–4 months were still observed[13].

Overall, we observed an average reduction of approximately 1.9 years in age at menarche over seven decades (i.e., an average reduction of approximately 0.1 years, or 36 days, per 5-year birth cohort, or roughly 72 days per decade). This decline is consistent with two previous Spanish studies conducted in other regions that reported decreases of approximately 36.5 days per 5-year birth cohort between 1940 and 1980[25] and 48 days per 5-year birth cohort between 1925 and 1962[24]. There is no comparable Spanish data from the 1980s onwards. Similar trends in other Western countries align with our findings. A study across nine European countries reported a decline of ∼44 days per 5-year birth cohort between 1918 and 1964. Country-specific estimates were lowest in the UK (18 days per 5-year birth cohort) and highest in Spain and Germany (nearly 58 days per 5-year birth cohort)[26]. Among single-country European studies, a Portuguese study reported estimates nearly identical to ours (decline of ∼31.1 days per 5-year birth cohort between 1920 and 2000)[40], whereas a Norwegian study found a less pronounced decrease (∼13 days per 5-year birth cohort among women born 1936–1964)[39]. Outside of Western countries, a large African study reported a mean overall secular decline of 29 days per 5-year birth cohort among women born 1935–1965, with steeper declines in sub-Saharan Africa and more modest or heterogeneous trends elsewhere[17]. In Latin America, the decline was markedly steeper in Colombia (born 1941–1989: ∼100 days per 5-year birth cohort)[43] than in Mexico (born <1940 to 1980: 27 days per 5-year birth cohort), the latter being more comparable with our results[44]. In Asia, studies conducted in China[45,46], Taiwan[16], Korea[47], India[15], and Indonesia[48] among women born in the mid- to late 20th century reported a secular decline of ∼0.035 to 0.36 years per 5-year birth cohort (∼13–130 days), generally larger than that observed in our study.

### 4.2. Determinants associated with the secular trend in menarche

Birth continent emerged as a key factor, with patterns differing depending on whether participants were classified as women or girls. Specifically, women born in South/Central America or Africa in the adult group (i.e., born ≤1975) tended to report a slightly later age at menarche compared to Europeans, whereas in the younger group (i.e., born from the 1990s onwards), those from South/Central America showed an earlier onset, while no data were available for girls from Africa. This reversed pattern among generations may reflect the complex interplay between biological factors and the socioeconomic and environmental context shaping pubertal development among migrant women[18,21]. Spain’s transition to a country of immigration since the 1990s[49] supports the interpretation that older foreign-born cohorts likely experienced menarche in their countries of birth, whereas younger cohorts may have migrated earlier in life and thus experienced menarche under Spanish nutritional and health conditions.

In our study, higher BMI in girls was associated with earlier menarche and lower BMI with later menarche. However, these analyses could not be conducted in women due to a lack of BMI data at menarche. Our findings are consistent with previous studies[45,47] and align with the well-established theory that adiposity is a key determinant of pubertal timing[20]. Biologically, higher BMI reflects greater body fat, which can accelerate pubertal onset via increased leptin, altered insulin sensitivity, and enhanced peripheral estrogen production[20]. Elevated estrogen can stimulate ovarian activity and the hypothalamic–pituitary–gonadal axis, promoting pubertal progression[20,50]. In this context, the increasing prevalence of overweight and obesity in children in Spain in recent decades[51] may partly contribute to the earlier menarche in our population. BMI is also closely related to lifestyle factors, such as diet and physical activity during childhood and adolescence[52], which may further influence pubertal timing.

Regarding education, the analyses were restricted to women because younger participants had not yet completed their education at the time of data collection. Associations were modest and did not show a clear gradient across categories. Only secondary education was statistically significantly associated with earlier menarche compared with primary education. The association between menarcheal age and education may reflect shared early-life socioeconomic conditions rather than causality, as education is often used as a proxy for socioeconomic status, yielding inconsistent results across studies. For example, studies in the United States[18] and United Kingdom[41] reported earlier menarche in lower socioeconomic groups, whereas Mexican[44] data showed the opposite pattern.

### 4.3. Limitations and strengths

Several limitations of this study should be acknowledged. First, information on social and environmental conditions was limited to continent of birth, BMI, and educational level, as only these variables were available in the two large databases, precluding the inclusion of additional variables from the smaller cohorts. In this context, beyond adiposity and sociodemographic factors, endocrine-disrupting chemicals with estrogenic activity may influence pubertal timing, as reported in the INMA cohort included in this study[53–56]. However, this information was unavailable in the large datasets and was not examined here. Second, limited sample sizes required the use of 5-year birth cohorts, and in some 5-year birth cohorts, insufficient numbers precluded the evaluation of determinants. Third, the long interval between menarche and reporting in adults may have reduced recall accuracy. Fourth, the assessment of the most recent cohort (born up to 2008) was limited because some girls had not yet reached menarche by 2019, potentially leading to an underestimation of more current trends. Fifth, the SIA-GAIA database recorded age at menarche as free text, which may have affected data completeness and consistency, potentially overrepresenting cases of precocious or delayed puberty.

Regarding the study’s strengths, first, the large, population-based sample (N=417,260), spanning 1931–2008, provides substantial statistical power and a unique opportunity to comprehensively assess long-term generational changes in pubertal timing within a defined socioeconomic and cultural context. Second, the use of multiple complementary data sources, including population-based registries and cohort studies, enhances the representativeness and validity of the findings. Third, the inclusion of women from diverse origins provides valuable insight into regional and ethnic differences in menarcheal age within the Valencian Community.

### 4.4. Conclusion

Our findings indicate a decrease in age at menarche among women in the Valencian Community born between 1931 and 2008, consistent with global evidence of a long-term downward trend. This decline was gradual among the earliest cohorts, followed by a temporary stabilization between 1960 and 1975, after which the decline resumed. The mean decrease was approximately 0.1 years (36 days) per 5-year birth cohort, equivalent to about 72 days per decade. Mean values dropped below 12 years in the youngest generations. Determinant analyses revealed the multifactorial nature of pubertal development, highlighting its sensitivity to nutritional, socioeconomic, and geographic factors. Notably, associations between continent of birth and age at menarche varied by birth cohort, with opposite patterns observed between adult and younger women born in South/Central America. This likely reflects differences in early-life conditions, with younger foreign-born cohorts raised in Spain reaching menarche earlier than older foreign-born cohorts who might have spent their childhood in their birth country. By contrast, higher BMI was consistently linked to earlier menarche, underscoring the importance of preventing childhood obesity for public health.

## Data availability statement

Data from population-based registries (Valencian Breast Cancer Screening Program and the Ambulatory Care Information System of the Valencian Health Agency) are not publicly available due to ethical and legal restrictions related to the use of sensitive health data, and access is restricted.

Data from the INMA and PAPILONG cohort studies are available upon request to the corresponding author and in accordance with the applicable data access procedures and ethical approvals. The INMA cohort data are subject to the INMA collaboration policy (https://www.proyectoinma.org/en/inma-project/inma-collaborationpolicy/).

## Acknowledgments

The authors gratefully acknowledge all participants for their generous collaboration, as well as the INMA consortium, the PAPILONG research team, the Valencian Breast Cancer Screening Program and the Ambulatory Care Information System of the Valencian Health Agency for their support and valuable contributions to this research.

## Funding

This article received funding from multiple sources. European Union funding was provided through the ATHLETE project (Grant Agreement No. 874583) under the Horizon 2020 programme, and the JAPreventNCD project (Grant Agreement No. 101128023), co-funded by the EU4Health Programme 2021–2027.

Additional funding was provided by Spanish institutions, including the Ministry of Science, Innovation and Universities (Grant CNS2023-145286, funded by MICIU/AEI/10.13039/501100011033 and by the European Union NextGenerationEU/PRTR); the Spanish Association Against Cancer (MICROVAGIPAP: IDEAS19098LOPE); the CIBER of Epidemiology and Public Health (PAPILONGO: ESP21PI03); the General Council of Official Nursing Associations of Spain (PAPISEX: inv_cge_2022_04); the Foundation for the Promotion of Health and Biomedical Research of the Valencian Region (PAPILONG: UGP-20-242); the Regional Ministry of Innovation, Universities, Science and Digital Society of the Generalitat Valenciana (MENTABIOTA: AICO/2021/182 and CIAICO/2023/184); and the Fundació La Marató (202414).

In addition, this article was supported by funding for research positions from the Carlos III Health Institute (Miguel Servet-FEDER: CP11/00178, MSII16/00051, CP20/0006, and Sara Borrell: CD23/00090, all co-funded by the European Union), as well as the Regional Ministry of Innovation, Universities, Science and Digital Society (Investigo contracts: INVEST/2022/310 and CIACIF/2022/268). The latter corresponds to a grant from the Programme for the Promotion of Scientific Research, Technological Development and Innovation in the Valencian Community, supporting the recruitment of predoctoral research staff and eligibility for co-financing by the European Social Fund.

## Author contributions

**Andrea Beneito:** Conceptualization; data curation; formal analysis; investigation; methodology; writing – original draft. **Blanca Sarzo:** Data curation; formal analysis; supervision; methodology; writing – original draft; writing – review and editing. **Raúl Beneyto:** Data curation; formal analysis; methodology; writing – original draft; writing – review and editing. **Reem Abumallouh, Natalia Marin, Oihane Alvarez, and Carmen Freire:** Data curation; funding acquisition; writing – review and editing. **Ana Molina-Barceló and Mercedes Vanaclocha-Espí:** Data curation; writing – review and editing. **Ferran Ballester:** Supervision; methodology; writing – review and editing. **Ana Esplugues:** Conceptualization; methodology; writing – review and editing. **Maria-Jose Lopez-Espinosa:** Conceptualization; data curation; formal analysis; funding acquisition; investigation; methodology; supervision; writing – original draft; writing – review and editing.

## Conflict of interest

No financial or non-financial benefits have been received or will be received from any party related directly or indirectly to the subject of this article.

## Notes

### Competing Interest Statement

The authors have declared no competing interest.

### Author Declarations

All study protocols, including two secondary population-based registries and two datasets collected by the research team, were approved by the Directorate of Public Health and the Higher Centre for Research in Public Health (DGSP-CSISP) Ethics Committee from Valencia (references numbers: 20180112/03; 20190329/3; 20190301/09/1; and 20210604/10/01). Secondary data were provided anonymized, while investigator-collected data were anonymized prior to analysis following written informed consent from participants or legal guardians (for minors). All procedures were conducted in accordance with the Declaration of Helsinki.

