## Supplementary material for "Secular trends in age at menarche and associated determinants in the Valencian population (Spain)"

Andrea Beneito, Blanca Sarzo, Raul Beneyto, [Reem Abumallouh](https://pubmed.ncbi.nlm.nih.gov/?term=Abumallouh+R&cauthor_id=35964788), Natalia Marin, Oihane Alvarez, Ana Molina-Barcelo, Mercedes Vanaclocha-Espi, Carmen Freire, Ferran Ballester, Ana Esplugues, Maria-Jose Lopez-Espinosa

**Table S1. Reported country of birth per continent**

| **Reported country** | **Continent** | **Women (N)** | | **Girls (N)** |
| --- | --- | --- | --- | --- |
|  | **Europe** |  | |  |
| Albania |  | 3 | | 0 |
| Andorra |  | 8 | | 0 |
| Austria |  | 34 | | 1 |
| Belarus |  | 21 | | 0 |
| Belgium |  | 490 | | 6 |
| Bosnia-Herzegovina |  | 17 | | 0 |
| Bulgaria |  | 1,178 | | 46 |
| Czechoslovakia |  | 13 | | 0 |
| Czech Republic |  | 27 | | 1 |
| Croatia |  | 11 | | 0 |
| Denmark |  | 31 | | 0 |
| Estonia |  | 11 | | 1 |
| Finland |  | 39 | | 0 |
| France |  | 4,475 | | 8 |
| Germany |  | 1,311 | | 9 |
| Greece |  | 4 | | 1 |
| Hungary |  | 40 | | 0 |
| Iceland |  | 5 | | 0 |
| Ireland |  | 88 | | 1 |
| Italy |  | 286 | | 20 |
| Latvia |  | 35 | | 2 |
| Lithuania |  | 235 | | 7 |
| Luxembourg |  | 12 | | 0 |
| Malta |  | 4 | | 0 |
| Moldova |  | 167 | | 2 |
| Monaco |  | 7 | | 0 |
| Montenegro |  | 1 | | 0 |
| The Netherlands |  | 427 | | 5 |
| North Macedonia |  | 3 | | 0 |
| Norway |  | 106 | | 4 |
| Poland |  | 244 | | 11 |
| Portugal |  | 243 | | 3 |
| Romania |  | 4,176 | | 133 |
| Russia |  | 706 | | 12 |
| Russian Federation |  | 5 | | 0 |
| Serbia |  | 2 | | 0 |
| Slovakia |  | 25 | | 0 |
| Slovenia |  | 11 | | 0 |
| Spain |  | 163,711 | | 11,704 |
| Sweden |  | 101 | | 1 |
| Switzerland |  | 469 | | 4 |
| United Kingdom |  | 3,053 | | 60 |
| Ukraine |  | 1,029 | | 28 |
| Vatican City |  | 4 | | 0 |
| Yugoslavia |  | 16 | | 5 |
| Total |  | 182,884 | | 12,075 |
|  | **South/Central America** |  |  | |
| Argentina |  | 1,820 | | 68 |
| Aruba |  | 1 | | 0 |
| Bahamas |  | 2 | | 0 |
| Bolivia |  | 1,035 | | 140 |
| Brazil |  | 632 | | 16 |
| Chile |  | 333 | | 14 |
| Colombia |  | 3,684 | | 153 |
| Costa Rica |  | 15 | | 0 |
| Cuba |  | 661 | | 12 |
| Dominica |  | 5 | | 0 |
| Dominican Republic |  | 341 | | 9 |
| Ecuador |  | 2,654 | | 131 |
| El Salvador |  | 27 | | 2 |
| Grenada |  | 1 | | 0 |
| Guatemala |  | 17 | | 0 |
| Honduras |  | 158 | | 18 |
| Jamaica |  | 3 | | 0 |
| Mexico |  | 160 | | 3 |
| Netherlands Antilles |  | 3 | | 0 |
| Nicaragua |  | 37 | | 3 |
| Panama |  | 14 | | 2 |
| Paraguay |  | 272 | | 13 |
| Peru |  | 524 | | 9 |
| Puerto Rico |  | 4 | | 0 |
| South America (unspecified) |  | 1 | | 0 |
| Uruguay |  | 740 | | 30 |
| Venezuela |  | 762 | | 49 |
| Total |  | 14,070 | | 685 |
|  | **Africa** |  | |  |
| Africa |  | 11 | | 0 |
| Algeria |  | 408 | | 5 |
| Angola |  | 9 | | 0 |
| Arabian Peninsula countries |  | 2 | | 0 |
| Benin |  | 1 | | 0 |
| Burkina Faso |  | 1 | | 1 |
| Cameroon |  | 11 | | 3 |
| Central African Republic |  | 1 | | 0 |
| Chad |  | 1 | | 0 |
| Congo |  | 12 | | 0 |
| Cape Verde |  | 18 | | 0 |
| Egypt |  | 5 | | 1 |
| Ethiopia |  | 5 | | 2 |
| Equatorial Guinea |  | 121 | | 15 |
| Gambia |  | 1 | | 0 |
| Ghana |  | 2 | | 2 |
| Guinea |  | 35 | | 1 |
| Guinea-Bissau |  | 1 | | 0 |
| Ivory Coast |  | 5 | | 0 |
| Kenya |  | 2 | | 0 |
| Lesotho |  | 1 | | 0 |
| Liberia |  | 2 | | 0 |
| Libya |  | 2 | | 0 |
| Mali |  | 3 | | 0 |
| Mauritius |  | 2 | | 0 |
| Mauritania |  | 6 | | 1 |
| Morocco |  | 2,227 | | 51 |
| Mozambique |  | 4 | | 0 |
| Nigeria |  | 24 | | 6 |
| Rhodesia |  | 1 | | 0 |
| São Tomé and Príncipe |  | 1 | | 0 |
| Senegal |  | 50 | | 2 |
| Sierra Leone |  | 1 | | 0 |
| South Africa |  | 15 | | 0 |
| Swaziland |  | 1 | | 0 |
| Tanzania |  | 2 | | 0 |
| Tunisia |  | 13 | | 1 |
| Zaire |  | 9 | | 0 |
| Zimbabwe |  | 2 | | 0 |
| Total |  | 3,009 | | 92 |
|  | **Asia** |  | |  |
| Afghanistan |  | 3 | | 0 |
| Armenia |  | 195 | | 14 |
| Azerbaijan |  | 7 | | 0 |
| Bahrain |  | 1 | | 0 |
| Bangladesh |  | 1 | | 0 |
| Myanmar |  | 1 | | 0 |
| Brunei |  | 2 | | 0 |
| Cambodia |  | 1 | | 0 |
| China |  | 448 | | 33 |
| Georgia |  | 71 | | 0 |
| India |  | 35 | | 3 |
| Indonesia |  | 7 | | 0 |
| Iraq |  | 5 | | 0 |
| Iran |  | 24 | | 1 |
| Japan |  | 24 | | 0 |
| Jordan |  | 3 | | 0 |
| Kazakhstan |  | 10 | | 1 |
| Kuwait |  | 1 | | 0 |
| Kyrgyzstan |  | 3 | | 0 |
| Laos |  | 4 | | 0 |
| Lebanon |  | 9 | | 0 |
| Malaysia |  | 7 | | 0 |
| Mongolia |  | 1 | | 0 |
| Nepal |  | 4 | | 0 |
| North Korea |  | 4 | | 0 |
| Pakistan |  | 65 | | 12 |
| Palestine |  | 1 | | 0 |
| Philippines |  | 78 | | 0 |
| Saudi Arabia |  | 1 | | 1 |
| Singapore |  | 3 | | 0 |
| South Korea |  | 4 | | 0 |
| South Vietnam |  | 1 | | 0 |
| Sri Lanka |  | 1 | | 0 |
| Syria |  | 28 | | 5 |
| Taiwan |  | 5 | | 0 |
| Thailand |  | 15 | | 0 |
| Turkey |  | 4 | | 0 |
| United Arab Emirates |  | 0 | | 1 |
| Uzbekistan |  | 5 | | 0 |
| Vietnam |  | 8 | | 0 |
| Total |  | 1,094 | | 71 |
|  | **North America & Oceania** |  |  | |
| Australia |  | 26 | | 0 |
| Canada |  | 30 | | 1 |
| North America |  | 3 | | 0 |
| United States of America |  | 152 | | 4 |
| Total |  | 211 | | 5 |

Note: N=sample size.

Participants who reported African or North American origin without specifying country of origin were grouped in the ‘Africa’ (N=11) or ‘North America’ (N=3) category. North America and Oceania were combined due to low numbers.

**Table S2. Number of participants per year of birth**

| **Year** | **1931** | **1932** | **1933** | **1934** | **1935** | **1936** | **1937** | **1938** | **1939** | **1940** |
| --- | --- | --- | --- | --- | --- | --- | --- | --- | --- | --- |
| **N** | 92 | 413 | 1,251 | 2,358 | 3,712 | 3,404 | 1,518 | 1,401 | 1,283 | 2,253 |
| **Year** | **1941** | **1942** | **1943** | **1944** | **1945** | **1946** | **1947** | **1948** | **1949** | **1950** |
| **N** | 1,855 | 2,174 | 2,579 | 2,742 | 2,973 | 3,061 | 3,438 | 3,900 | 4,020 | 4,394 |
| **Year** | **1951** | **1952** | **1953** | **1954** | **1955** | **1956** | **1957** | **1958** | **1959** | **1960** |
| **N** | 4,852 | 8,118 | 14,716 | 18,637 | 19,486 | 19,425 | 20,044 | 21,473 | 21,414 | 22,277 |
| **Year** | **1961** | **1962** | **1963** | **1964** | **1965** | **1966** | **1967** | **1968** | **1969** | **1970** |
| **N** | 21,733 | 18,672 | 16,164 | 17,185 | 14,921 | 21,926 | 12,659 | 11,783 | 9,811 | 10,382 |
| **Year** | **1971** | **1972** | **1973** | **1974** | **1975** | **1976** | **1977** | **1978** | **1979** | **1980** |
| **N** | 7,820 | 8,537 | 5,667 | 109 | 30 | 22 | 22 | 15 | 4 | 10 |
| **Year** | **1981** | **1982** | **1983** | **1984** | **1985** | **1986** | **1987** | **1988** | **1989** | **1990** |
| **N** | 3 | 7 | 1 | 0 | 1 | 0 | 0 | 0 | 0 | 51 |
| **Year** | **1991** | **1992** | **1993** | **1994** | **1995** | **1996** | **1997** | **1998** | **1999** | **2000** |
| **N** | 229 | 454 | 758 | 1,129 | 1,196 | 1,386 | 1,403 | 1,421 | 1,595 | 1,589 |
| **Year** | **2001** | **2002** | **2003** | **2004** | **2005** | **2006** | **2007** | **2008** |  |  |
| **N** | 1,539 | 1,531 | 1,553 | 1,637 | 1,589 | 1,036 | 405 | 12 |  |  |

Note: **N**=sample size

**Table S3. Descriptive statistics by 5-year birth cohort**

|  |  | **Population included in the temporal series** | |  | **Population included in the Bayesian analyses^a^** | | |
| --- | --- | --- | --- | --- | --- | --- | --- |
| **5-year birth cohort** | **Sample size** | **Age at menarche (years)** | |  | **Sample size** | **Age at menarche (years)** | |
|  | **N** | **Mean ± SD** | **Median (P25, P75)** |  | **N** | **Mean ± SD** | **Median (P25, P75)** |
|  | **Women (N=396,747)** | | |  | **Women (N=177,025)** | | |
| **1931–1935** | 7,826 | 13.1±1.65 | 13 (12, 14) |  | 0 | - | - |
| **1936–1940** | 9,859 | 12.9±1.67 | 13 (12, 14) |  | 0 | - | - |
| **1941–1945** | 12,323 | 12.7±1.64 | 13 (12, 14) |  | 472 | 12.8±1.57 | 13 (12, 14) |
| **1946–1950** | 18,813 | 12.6±1.59 | 13 (12, 14) |  | 2,099 | 12.8±1.61 | 13 (12, 14) |
| **1951–1955** | 65,809 | 12.6±1.55 | 13 (11, 14) |  | 13,037 | 12.6±1.58 | 13 (12, 14) |
| **1956–1960** | 104,633 | 12.5±1.56 | 13 (11, 14) |  | 22,759 | 12.6±1.58 | 13 (11, 14) |
| **1961–1965** | 88,675 | 12.7±1.56 | 13 (12, 14) |  | 66,014 | 12.6±1.56 | 13 (12, 14) |
| **1966–1970** | 66,561 | 12.7±1.58 | 13 (12, 14) |  | 56,677 | 12.7±1.57 | 13 (12, 14) |
| **1971–1975** | 22,163 | 12.7±1.59 | 13 (12, 14) |  | 15,967 | 12.7±1.59 | 13 (12, 14) |
| **1976–1980** | 73 | 12.3±1.70 | 13 (11, 13) |  | 0 | - | - |
| **1981–1985** | 12 | 12.3±1.60 | 12 (11, 13) |  | 0 | - | - |
|  | **Girls (N=20,513)** | | |  | **Girls (N=11,867)** | | |
| **1986–1990^b^** | 51 | 12.2±1.57 | 12 (11, 13) |  | 0 | - | - |
| **1991–1995** | 3,766 | 12.0±1.35 | 12 (11, 13) |  | 2,710 | 12.0±1.34 | 12 (11, 13) |
| **1996–2000** | 7,394 | 11.7±1.18 | 12 (11, 13) |  | 5,463 | 11.7±1.18 | 12 (11, 12) |
| **2001–2005** | 7,849 | 11.6±1.10 | 12 (11, 12) |  | 3,694 | 11.6±1.10 | 12 (11, 12) |
| **2006–2008^c^** | 1,453 | 11.1±0.84 | 11 (11, 12) |  | 0 | - | - |

Note: ^a^Bayesian analyses of determinants associated with the secular trend of menarche were conducted in a subsample, including only participants with complete data on all the studied determinants and excluding birth cohorts 1931–1940, 1976–1990, and 2006–2008 (the last truncated at 2008 due to unavailable data) because of small sample sizes with determinant information, and excluding participants born in North America, Oceania, Asia (analyses of women and girls) and Africa (analyses of girls) due to small sample sizes. **^b^**For the 1986–1990 birth cohort, all girls were born in 1990. ^c^For the most recent 5-year birth cohort (2006–2010), data availability was limited to births up to 2008.

Age at menarche is presented as mean ± standard deviation (SD) and median (25th–75th percentiles, P25–P75).

**Table S4. Predicted value and 95% confidence interval (95% CI) of age at menarche for the next 5-year birth cohorts**

| **5-year birth cohort: range** | **Predicted age at menarche (95% CI): years** |
| --- | --- |
| 2011–2015 | 10.8 (10.5, 11.1) |
| 2016–2020 | 10.5 (10.0, 11.1) |
| 2021–2025 | 10.2 (9.42, 11.0) |
| 2026–2030 | 9.89 (8.82, 11.0) |
| 2031–2035 | 9.58 (8.20, 11.0) |

Note: CI: confidence interval

**Table S5. Descriptive characteristics of the population included in the Bayesian analyses by age group**

| **Variable** | **Overall**  **(N=188,892)** | **Women**  **(N=177,025)** | **Girls**  **(N=11,867)** |
| --- | --- | --- | --- |
| Age at menarche (years) | 12.6±1.57 | 12.6±1.57 | 11.8±1.20 |
| Continent of birth: n (%) |  |  |  |
| Europe | 174,622 (92.4) | 163,369 (92.3) | 11,253 (94.8) |
| South/Central America | 11,945 (6.32) | 11,331 (6.40) | 614 (5.17) |
| Africa | 2,325 (1.23) | 2,325 (1.31) | – |
| BMI (z-score) |  |  |  |
| Low BMI | – | – | 110 (0.93) |
| Normal BMI | – | – | 6,683 (56.3) |
| High BMI | – | – | 5,074 (42.8) |
| Educational level: N (%) |  |  |  |
| Primary education | – | 92,825 (52.4) | – |
| Secondary education | – | 51,090 (28.9) | – |
| University education | – | 33,110 (18.7) | – |

Note: “–” indicates absence of observations in the corresponding category. BMI: body mass index based on WHO age- and sex-specific z-scores[1] and classified as low (≤–2 standard deviations [SD]), normal (>–2 to ≤1 SD), or high (>1 SD).

Categorical variables are shown as N (%) and continuous variables as mean ± SD. Subsample derived from the total study population described in Table 1, restricted to participants with complete data on the studied determinants and used in the Bayesian analyses. Educational level and BMI were available only for women and girls, respectively. The number of girls born in Africa was low and therefore not included in the Bayesian analyses; however, corresponding data are presented here for the overall population.

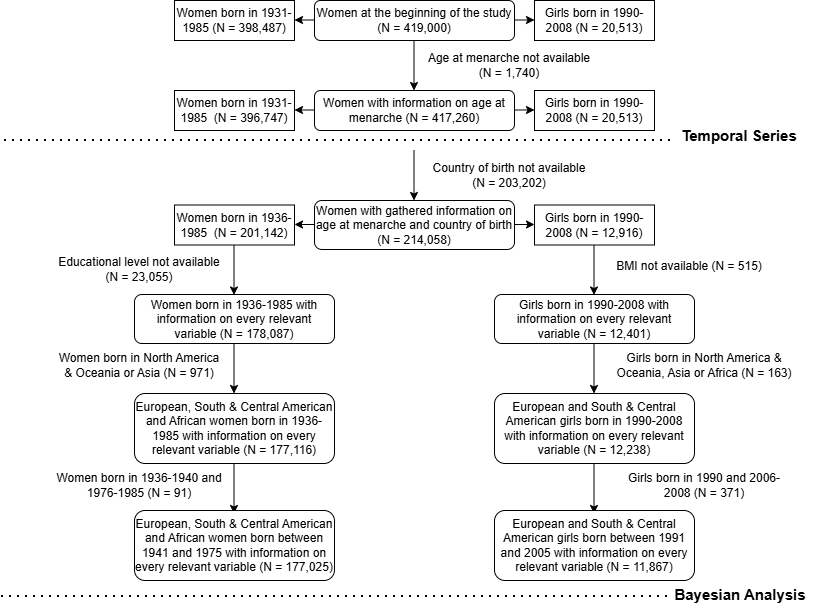

**Figure S1. Flowchart showing the evolution from the initial to the final population sample used in this study**

Two populations were used in the present study: one for the temporal series and another for the Bayesian analyses. Bayesian analyses of the determinants associated with the secular trend of menarche were conducted in a subsample, including only participants with complete data on all the studied determinants and excluding birth cohorts 1931–1940, 1976–1990, and 2006–2008 (the last truncated at 2008 due to unavailable data) because of small sample sizes with determinant information, and excluding participants born in North America, Oceania, Asia (analyses of women and girls), and Africa (analyses of girls) due to small sample sizes. Separate Bayesian linear regression models were fitted for each group, adjusting for birth cohort and continent of birth in all models, and additionally for educational level in women and body mass index (BMI) in girls.

For the 1986–1990 birth cohort, all girls were born in 1990. For the most recent 5-year birth cohort (2006–2010), data availability was limited to births up to 2008.

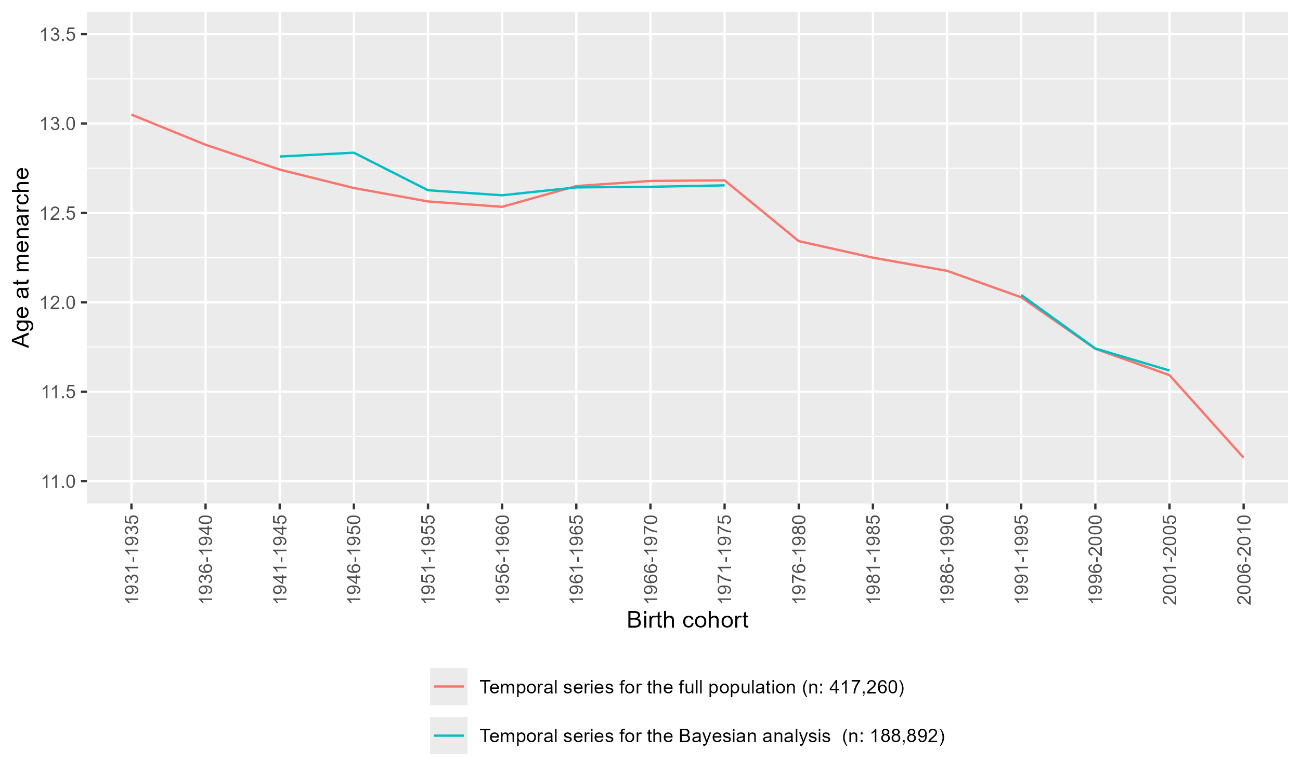

**Figure S2. Sensitivity analysis: Comparison of secular trend in age at menarche (years) between the initial population (temporal series analysis; red line) and the population included in the Bayesian analysis (blue line)**

Note: N=sample size. Red line: women and girls included in the temporal series analysis born between 1931 and 2008. Blue line: population included in the Bayesian analysis, women born between 1941 and 1975 and girls born between 1991 and 2005. For the most recent 5-year birth cohort (2006–2010), data availability was limited to births up to 2008.

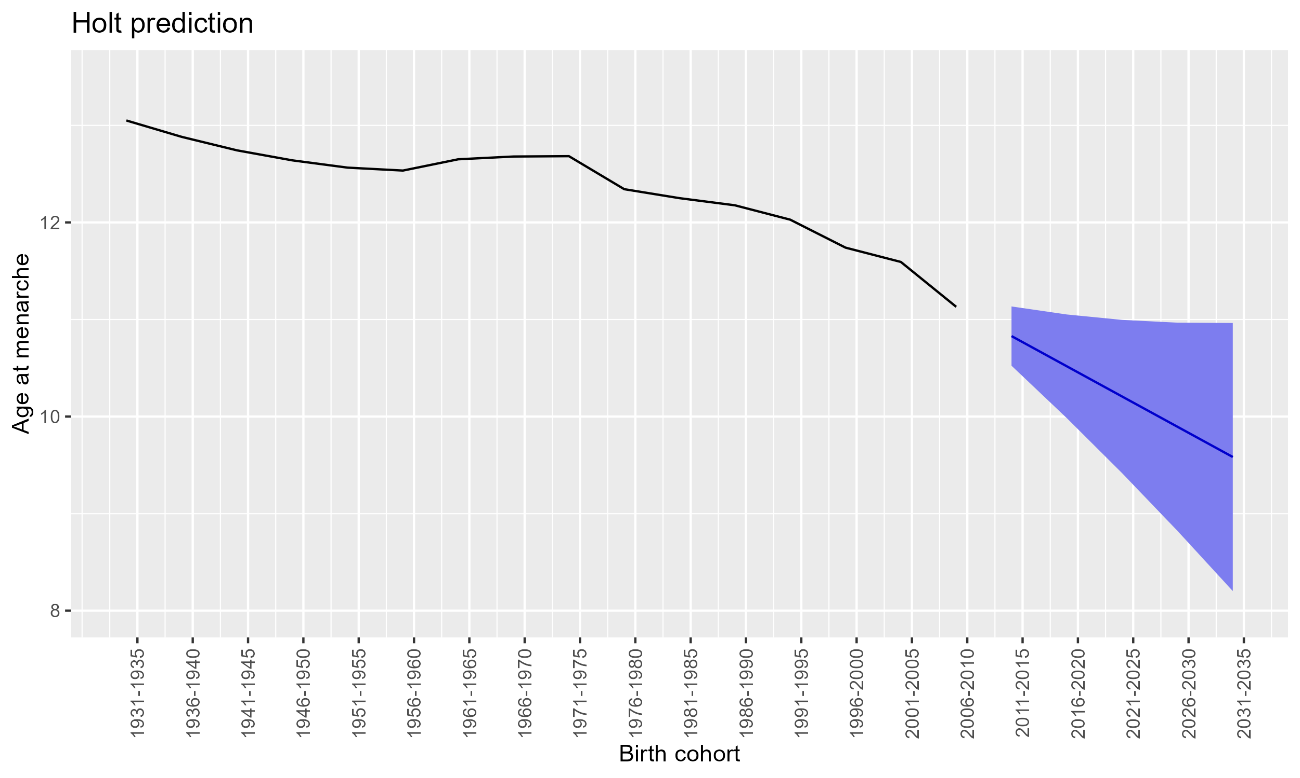

**Figure S3. Temporal series analysis of age at menarche (years), including women born between 1931 and 2008, with predicted data (in blue) for the years between 2011 and 2035**

Note: For the most recent 5-year birth cohort (2006–2010), data availability was limited to births up to 2008.
